# Baseline resting EEG measures differentiate rTMS treatment responders and non-responders

**DOI:** 10.1101/2023.11.16.23298445

**Authors:** Pakin Kaewpijit, Paul B Fitzgerald, Kate Hoy, Neil W Bailey

## Abstract

**Background:** Repetitive transcranial magnetic stimulation (rTMS) has been increasingly used worldwide in the treatment of depression, however, we currently lack the means to reliably predict whether patients will respond to the treatment. Recent research suggests that the neurophysiological measures of beta power and correlation dimension may have predictive potential, however, studies of beta power and correlation dimension to differentiate rTMS group response in individuals with major depression are limited.

**Methods:** Fifty treatment-resistant patients with major depressive disorder were recruited. Forty-two participants underwent baseline resting EEG sessions and 5-8 weeks of rTMS treatments and 12 participants were responders to the treatment. Beta power and correlation dimension from baseline resting EEG were compared between responders and non-responders.

**Results:** Responders demonstrated significantly lower beta power in baseline resting EEG, however, correlation dimension did not show a significant difference between groups.

**Limitations:** There were a small number of responders in this study.

**Conclusion:** Baseline resting beta power may help to differentiate responders from non-responders to rTMS treatment. However, further studies are needed with larger sample sizes.

## Introduction

Major depressive disorder (MDD) is a common psychiatric illness. Approximately 30% of patients with MDD do not respond to standard medication and psychological therapies^1^. Repetitive transcranial magnetic stimulation (rTMS) is an increasingly available and effective treatment but not all participants respond, with studies reporting a wide range of response and remission rates, 29-58% and 6-37%, respectively^2-4^. Due to the high cost and time-consuming nature of the standard rTMS protocol, an effective treatment-response predictor would help physicians make a more well-informed decision about whether to recommend rTMS treatment.

Many studies have attempted to find demographic and clinical variables that provide accurate prediction of treatment response, but to date, no studies have found a consistently (replicable) strong relationship to differentiate responders from non-responders^5^. Functional magnetic resonance imaging (fMRI) and electroencephalography (EEG) have been widely used for studying the pathophysiology and neural circuitry of depression and for testing potential treatment-response predictors. One fMRI study has shown that one of four subgroups categorised by pre-treatment functional connectivity had a greater response to rTMS treatment provided to the dorsomedial prefrontal area^6^. Unfortunately, flaws have been suggested in their statistical testing method, and a recent non-replication of their results has been reported^7^. Research has also indicated that stimulating a DLPFC target that is functionally negatively correlated with subgenual anterior cingulate cortex (sgACC) is the most effective target for depression symptoms reduction^8^. Moreover, a greater clinical response was associated with higher baseline sgACC metabolism and decreased sgACC metabolism after treatment^9^. These results are promising for the prediction of response to rTMS for depression, however, due to its cost, fMRI is not extensively available, especially in the clinical setting.

EEG is another measure that can potentially be used to predict treatment response. For the prediction of response to treatment for MDD, many types of EEG measures have been examined, including resting EEG, sleep EEG, event-related potentials (ERPs) and transcranial magnetic stimulation-evoked EEG (TMS-EEG)^10-12^. Some EEG measures have been investigated both to help diagnose MDD and predict treatment response from both pharmacological treatment and other therapeutic interventions^13^. There have been resting EEG measures examined such as peak alpha frequency proximity to the stimulation frequency, the bispectrum of all frequency bands, and also nonlinear features (i.e., correlation dimension (CD), Lempel-Ziv complexity and Katz fractal dimension (KFD)) which have shown predictive potential^14,15^. The relationship between peak alpha frequency proximity to the stimulation frequency and treatment response has been shown to predict response to rTMS treatment for depression^15^, a finding that has been replicated in an independent dataset, suggesting that it might reflect a generalisable predictor^16^. If other potential predictors can be demonstrated to replicate, a combination of these generalisable predictors might provide accurate and clinically applicable treatment response predictions. This provides strong justification for replication studies of potential treatment response predictors.

Beta oscillations are high-frequency brain activity ranged from 13-30 Hz. They can be seen in an awake state, especially when subjects are anxious, stressed or highly arousal. In the frontocentral regions, beta oscillations usually have maximal amplitude^17,18^. Higher beta activity has also been found in patients with a recurrent depression^19^. Another measure that was also shown to provide predictive accuracy is the correlation dimension of the EEG, which is a way to measure the dimensionality of the data, which represents the trajectory of dynamical system. When the correlation dimension is low, there is low complexity in the system^20,21^. In EEG analysis, the correlation dimension was first used to study human brain activity while sleeping^22^. It was found that there was less dimensionality in deeper sleep stages. The correlation dimension has also been used to differentiate people with and without depression with high accuracy compared to other non-linear features (i.e., detrended fluctuation analysis, Higuchi fractal and Lyapunov exponent)^23^. Interestingly, two recent studies using machine learning techniques found that beta power and correlation dimension accurately predicted clinical response to rTMS^14,24^, holding promise for potential clinical application. However, robust determination of an effect that might be used clinically requires replication in independent datasets. As such, it is important to test if the effect replicates in other datasets. As such, we aimed to conduct an exploratory analysis of potential beta power and correlation dimension differences between responders and non-responders in an independent dataset, in order to confirm the findings of previous research. The objective of this study was to examine whether beta power and correlation dimension can differentiate responders to rTMS treatment from non-responders.

## Methods

### Participants

Participants included in the current study were identical to those reported in our previous research on resting EEG predictors of response to rTMS treatment for depression^25^, which was part of a larger treatment study to assess the effect of switching rTMS treatment approach in the event of non-response^26^. Here we report on data only from the MDD participants and only from the baseline EEG. We recruited 50 major depressive disorder participants. Participants were aged between 20 and 72 years. Inclusion criteria for the MDD group were a DSM-IV diagnosis of major depressive episode using the MINI V5.0.0^27^, aged between 18–75, treatment resistant MDD at stage 2 of the Thase and Rush (1997) classification, a Montgomery-Asberg Depression Rating Scale (MADRS) score of >20, and no change to medication in the four weeks prior to screening or for the duration of the trial. As is usual in rTMS trials for treatment resistant depression, a number of participants in the MDD group were medicated^26^. Exclusion criteria were: presence of an unstable medical condition or neurological disorder; history of seizure; contraindication to rTMS; or current pregnancy or lactating. Participants were also excluded if at sufficient risk of suicide to require immediate electroconvulsive therapy, or had a current DSM-IV diagnosis of substance abuse or dependence disorder, bipolar affective disorder, or schizophrenia spectrum disorders. Ethical approval was granted by the Alfred Hospital and Monash University’s ethics committees, and all participants gave written informed consent.

Relevant to the current analyses, all participants had a baseline EEG session which was conducted prior to their first treatment session. Eight MDD participants were excluded from analysis because they withdrew from the study before the end of the first week of rTMS treatment. For the purposes of the current analyses of the remaining 42 MDD participants, were defined 12 as responders using a criteria of by >50% reduction in the 17-item Hamilton Rating Scale for Depression (HAM-D), as per our previous research on measures that differentiated responders and non-responders in this dataset^28,29^.

### Procedure

The demographic and depression severity data were collected at baseline interview. We used the 17-item Hamilton Rating Scale for Depression (HAM-D)^28^, the Montgomery-Asberg depression rating scale (MADRS)^30^ and the Beck Depression Inventory-II (BDI-II)^31^ for assessing depression severity. MDD participants underwent unilateral left 10 Hz rTMS treatment for three weeks (five days per week). Responders at the end of the third week continued to receive an additional two weeks of titrated rTMS treatment (three sessions in week four and two sessions in week five). Non-responders at three weeks (defined as less than a 25% reduction in their MADRS scores at this three week timepoint) were randomised to either continue for the next three weeks with unilateral left 10 Hz rTMS treatment, switch to unilateral right 1 Hz rTMS, or switch to sequential bilateral rTMS consisting of right 1 Hz rTMS followed by left 10 Hz rTMS treatment. Individuals who had not responded by week three, but responded by week six were continued on an additional 2 weeks of titrated rTMS treatment using the treatment approach they had been switched to (three sessions in week seven and two sessions in week eight). All rTMS treatments were given at 110% of resting motor threshold. Left sided treatment consisted of 40 trains of 5 s durations with a 25 s inter-train interval. Right sided rTMS consisted of 1 train of 1200 pulses. Bilateral rTMS combined these protocols but with only 900 right sided pulses. Treatments were given to the F3 and F4 electrode locations. MADRS and BDI-II measures were repeated at the end of week one and week three. The MADRS, BDI-II, and HAM-D were measured at week six, and at week eight for individuals who had not responded by week three, but had responded by week six.

EEG recordings and pre-processing were identical to Bailey et al. (2019)^25^. All participants provided 63 or more noise free epochs of two seconds in length, which were available for analysis from eyes open and eyes closed resting recordings, providing enough trials for reliable analysis. Data were re-referenced to an averaged reference offline.

### Beta Power analysis

EEG signal from each accepted epoch for each participant were submitted to a multitaper fast Fourier frequency transformation with a Hanning taper in order to calculate power in the beta band (12–30 Hz). Average power was calculated across the entire epoch within each frequency band, and then averaged across all trials from eyes open and eyes closed conditions together, resulting in a single value for each participant at each electrode. Statistical comparisons were made between groups and time points using these participant averages.

### Correlation dimension analysis

Correlation dimension is a nonlinear measure that can be applied to EEG data to measure the complexity of EEG signals^32^. Average correlation dimension was calculated for each electrode for each epoch, then averaged across epochs resulting in a single value for each participant from each electrode in the eyes closed condition. Statistical comparison was made between groups using these participant averages.

### Primary Comparisons

Primary statistical comparisons between groups in beta power EEG data were performed using RAGU^33^. RAGU uses reference free root mean square (RMS) measures and randomisation statistics to compare neural response strength and scalp field differences across all electrodes without a priori assumptions about locations that show significant effects. RAGU controls for multiple comparisons in the spatial dimension by collapsing differences to a single scalp difference map value for comparisons of the distribution of activity across the scalp, and using the single RMS value from each participant for global neural activity strength comparisons. Further details about RAGU can be found in Koenig, Kottlow^33^. Differences between groups in overall neural activity strength within the beta frequencies (across all electrodes) were compared using the RMS test. Differences between groups in the distribution of activity across the scalp were compared independently of amplitude using the topographical analysis of variance (TANOVA) after the recommended L2 normalization (which normalizes for global neural activity strength by converting data from each participant to the same RMS value). RMS and TANOVA tests were used to conduct repeated measure ANOVA design comparisons comparing beta power values averaged across all beta frequencies between responders and non-responders and within eyes open and eyes closed conditions. Because EEG measure can be affected by age^34^, independent t-tests was used to compare mean age between responder and non-responder groups.

The primary comparison between groups for correlation dimension was a repeated measures ANOVA design, with 2 groups x 28 electrodes conducted in SPSS 26. Only the main effect of group or interaction between group and electrode was of interest.

## Results

There were no significant differences in the ages of the responder and non-responder groups t(40) = 0.4934, p = 0.6244.

### Beta power

A significant group difference was present in the RMS test for averaged beta power (p = 0.026, η2 = 0.114) such that responders to rTMS displayed lower BL beta power compared to non-responders. No interaction between group and eyes open/closed was present (p = 0.213, η2 = 0.045). No significant group difference was present in the TANOVA test for averaged beta power (p = 0.624, η2 = 0.019), nor was there a significant interaction between group and eyes open/closed (p = 0.903, η2 = 0.012), suggesting that no significant difference was present in the topographical distribution of beta activity between two groups.

**Fig. 1.**
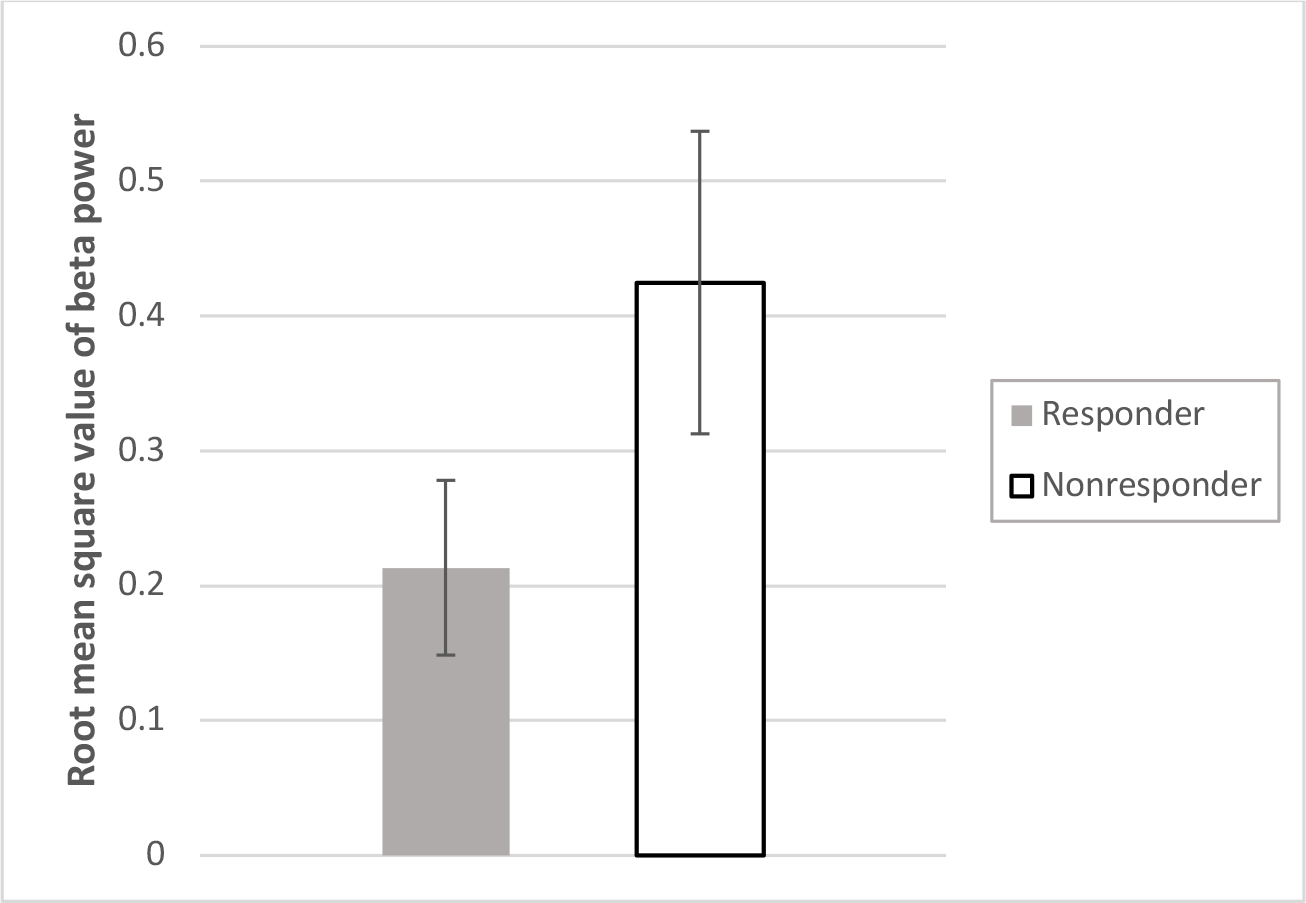
Mean root mean square value of beta power from responder and non-responder group. A significant difference was detected between responders and non-responders. Error bars reflect 95% confidence interval.

### Correlation dimension

There was no significant group difference in eyes-closed correlation dimension (F(1,40) = 0.570, p = 0.455, ηp2 = 0.014). Nor was there an interaction between group and electrode (F(1,40) = 1.303, p = 0.138, ηp2 = 0.032).

**Fig. 2.**
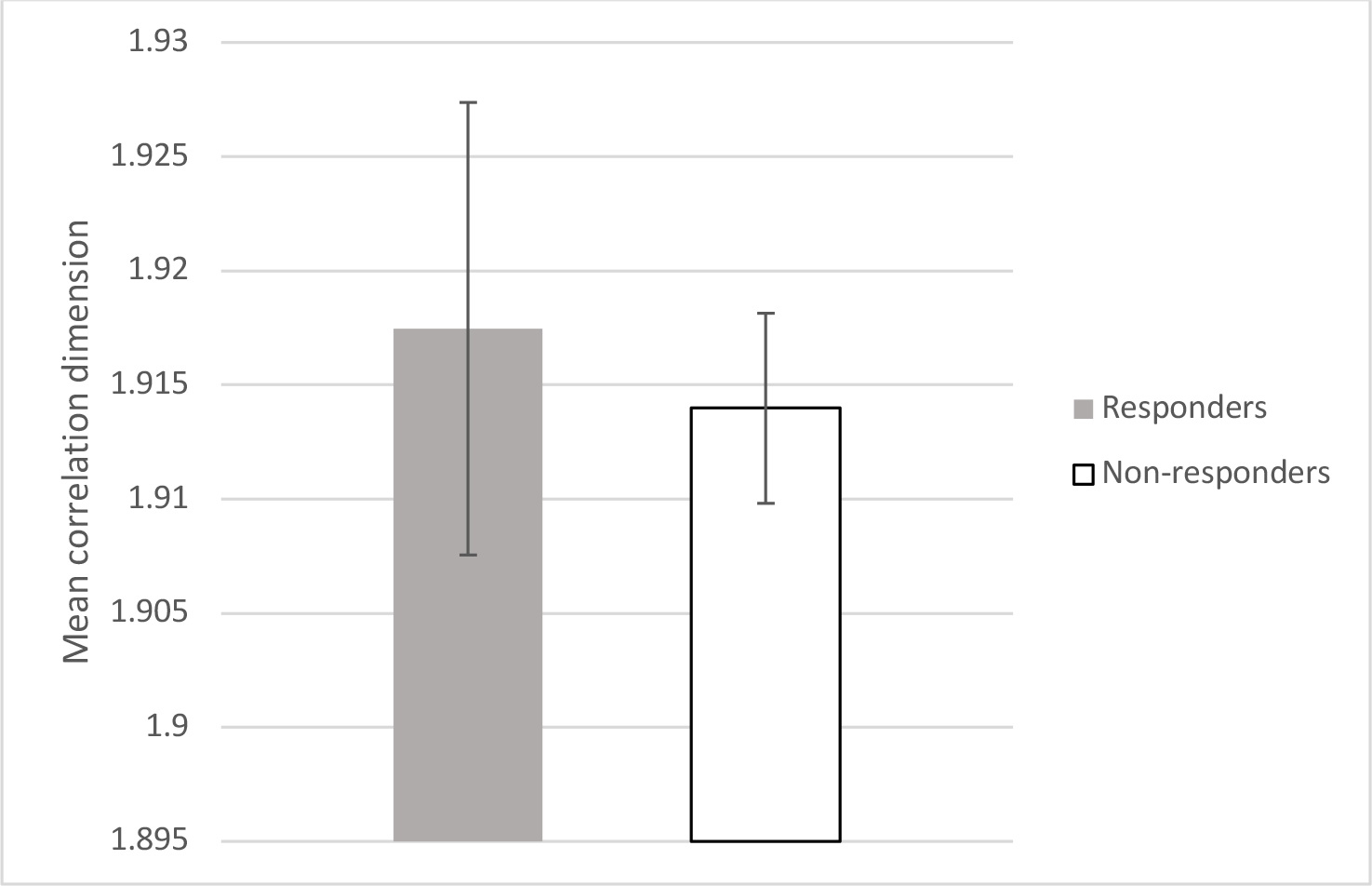
Mean correlation dimension values from responder and non-responder groups. No significant difference was detected. Error bars reflect 95% confidence interval.

## Discussion

Our study aimed to examine whether previously reported baseline beta power and correlation dimension measures are generalizable at differentiating rTMS treatment responders and non-responders. Our results indicated that responders to rTMS treatment had significantly less beta power than non-responders in baseline resting EEG recordings in both eyes-open and eyes-closed combined conditions. The result was similar to those reported by Hasanzadeh et al., which they reported high predictive accuracy of 91.3% for beta power^14^, and Ebrahimzadeh et al.^24^, who also reported that beta power predicted response to rTMS treatment for depression. However, our result extends their results by demonstrating that the differences in beta power are present in both the resting eyes-closed condition and the eyes open resting state (in contrast to the previous research which only examined the eyes closed state)^14^. There were differences in our rTMS treatment protocol compared to that tested by Hasanzadeh et al.’, but a similar number of rTMS sessions and similar treatment sites were applied. As such, our results indicate that beta power differentiates responders and non-responders to rTMS treatment for depression, indicating the finding replicates across multiple independent datasets, suggesting it may be a reliable and generalizable measure for differentiating responders and non-responders. This justifies further research using this measure, ideally using a prospective prediction study where participants are allocated to rTMS or another treatment based on their predicted responses. Interestingly, previous research has also indicated that both absolute and relative beta power are negatively associated with HAM-D score reduction in 6 weeks in pharmacological treatment (i.e., paroxetine)^35^. Similarly, another study demonstrated that higher beta power at the baseline resting EEG was correlated with more severe depressive symptoms after 4 week of antidepressant treatment^36^. Therefore it is possible that beta power may be not only useful to differentiate responders and non-responders to rTMS treatment, but also to antidepressant medications.

In contrast to our findings for beta power, our results showed no differences in correlation dimension between groups. This lack of replication of the research by Hasanzadeh et al., which they reported high predictive accuracy of 87% for correlation dimension^14^, could be explained by differences in concurrent medications, different complexity measures and participants’ age. However, if these differences explained the null result for the correlation dimension, it is not clear why the same would not be true for the beta finding. As such, it may be that the correlation dimension finding reported by Hasanzadeh et al., reflects a spurious finding. Alternatively, differences in study parameters, for example the EEG recording or pre-processing settings could also conceivably have affected our result, and it is possible these explain the difference between our studies. For example, there is evidence that correlation dimension increases with age in the healthy population because of the development and modification of neural cells and learning processes throughout lifespan^37^, so potential differences in the ages of participants between in our study and that have Hasanzadeh could be a potential explanation for the conflict in our results. Nevertheless, research in correlation dimension in depressive population is limited^38^ and needs further investigation.

### Limitations

There are some limitations in this study. Firstly, the sample size was not large and the number of responders was only 12 participants. This may offer a potential explaination for our non-significant correlation dimension result. Moreover, a small sample size and only a few measures in this study limited our capacity to conduct a valid machine learning analysis in direct replication of Hasanzadeh to determine prediction accuracy. Secondly, participants were taking many medications which affect EEG^39^, although we note that this factor is difficult to control for when conducting research in real clinical settings. Lastly, the rTMS protocol applied in this study was different to the FDA-approved rTMS protocol, as participants that showed signs of non-response at 3 weeks were re-randomised to either receive continued high-frequency left hemisphere rTMS treatment, or low-frequency right hemisphere rTMS treatment, or bilateral rTMS treatment. The difference of rTMS protocol and treatment stimulating sites across studies may affect EEG measures to an unknown degree so caution should be considered in interpreting our results.

## Conclusions

Global beta power differentiated responders to rTMS treatment for depression from non-responders from baseline EEG data, in replication of two previous studies. Correlation dimension was not significantly different between two groups. Further studies are needed with larger sample size, standard rTMS protocol applied and the consideration of EEG recording type and concurrent medications to determine if these results might be considered in a prospective prediction study and also in clinical application.

## Data Availability

All data produced in the present study are available upon reasonable request to the authors.

## Abbreviations

rTMS: repetitive transcranial magnetic stimulation
fMRI: functional magnetic resonance imaging
sgACC: subgenual anterior cingulate cortex
DLPFC: dorsolateral prefrontal cortex
BL: baseline
EEG: electroencephalography
MDD: major depressive disorder
HAM-D: Hamilton Rating Scale for Depression
BDI-II: Beck depression inventory
MADRS: Montgomery Asberg Depression Rating Scale
RMS: root mean square
TANOVA: topographical analysis of variance

