## Supplement materials for "Baseline resting EEG measures differentiate rTMS treatment responders and non-responders"

RAGU (Randomization Graphical User interface)

Ragu is a MATLAB-based open source toolbox for statistical analyses of multichannel EEG. When frequency comparisons are computed with RAGU, the average reference is not computed on the frequency transformed data (the average reference was computed prior to the frequency transforms). As such, the test is a comparison of the root mean square (RMS) between groups, a measure which is a valid indicator of neural response strength in the frequency domain^1^.
